# Rad-Path Correlation of Deep Learning Models for Prostate Cancer Detection on MRI

**DOI:** 10.1101/2025.06.04.25328868

**Authors:** A. S. C. Verde, J. G. de Almeida, F. Mendes, M. Pereira, R. Lopes, M. J. Brito, M. Urbano, P. S. Correia, A. M. Gaivão, A. Firpo-Betancourt, J. Fonseca, C. Matos, D. Regge, K. Marias, M. Tsiknakis, ProCAncer-I Consortium, R. C. Conceição, N. Papanikolaou

## Abstract

While Deep Learning (DL) models trained on Magnetic Resonance Imaging (MRI) have shown promise for prostate cancer detection, their lack of direct biological validation often undermines radiologists’ trust and hinders clinical adoption. Radiologic-histopathologic (rad-path) correlation has the potential to validate MRI-based lesion detection using digital histopathology. This study uses automated and manually annotated digital histopathology slides as a standard of reference to evaluate the spatial extent of lesion annotations derived from both radiologist interpretations and DL models previously trained on prostate bi-parametric MRI (bp-MRI). 117 histopathology slides were used as reference. Prospective patients with clinically significant prostate cancer performed a bp-MRI examination before undergoing a robotic radical prostatectomy, and each prostate specimen was sliced using a 3D-printed patient-specific mold to ensure a direct comparison between pre-operative imaging and histopathology slides. The histopathology slides and their corresponding T2-weighted MRI images were co-registered. We trained DL models for cancer detection on large retrospective datasets of T2-w MRI only, bp-MRI and histopathology images and did inference in a prospective patient cohort. We evaluated the spatial extent between detected lesions and between detected lesions and the histopathological and radiological ground-truth, using the Dice similarity coefficient (DSC). The DL models trained on digital histopathology tiles and MRI images demonstrated promising capabilities in lesion detection. A low overlap was observed between the lesion detection masks generated by the histopathology and bp-MRI models, with a DSC = 0.10. However, the overlap was equivalent (DSC = 0.08) between radiologist annotations and histopathology ground truth. A rad-path correlation pipeline was established in a prospective patient cohort with prostate cancer undergoing surgery. The correlation between rad-path DL models was low but comparable to the overlap between annotations. While DL models show promise in prostate cancer detection, challenges remain in integrating MRI-based predictions with histopathological findings.

## Introduction

The current gold standard for prostate cancer diagnosis in men with elevated PSA or associated risk factors is to perform a multi or bi-parametric magnetic resonance imaging (MRI) examination before biopsy ^1^. Early and precise lesion detection and characterization through MRI can help with patient selection for targeted biopsies to complement systematic sampling.

While Deep Learning (DL) models show promise in addressing the limitations of traditional MRI interpretations, such as inter- and intra-reader variability and the ability to detect “invisible lesions” ^2^, their translation into clinical practice is hindered by issues related to trustworthiness, generalizability, usability, and regulatory frameworks. These barriers underscore the need for further development and validation of DL models to ensure their reliability and effectiveness in real-world clinical settings. Conducting biological validation ^3^ through correlation with histopathology slides can improve trust in the outputs of these DL models. Since tissue biopsies are integral to prostate cancer diagnosis, correlating MRI with histopathological information is invaluable for mapping tumor extent, identifying previously undetected tumors, planning treatments, and validating artificial intelligence models. This form of biological validation is particularly promising in cases where whole tissue samples are accessible post-resection, such as those undergoing radical prostatectomy.

Previous studies have attempted to correlate prostate MRI with histopathology. For instance, Turkbey et al. ^4^ used a cohort of 45 prospective patients with 3D-printed patient-specific molds to correlate imaging with histopathology to validate MRI for prostate cancer detection. McGarry et al. ^5^ relied on aligned rad-path datasets of 39 patients to use unique image signatures to generate predictive maps of prostate cancer localization. McGarry et al. ^6^ used image signatures from 48 patients to predict histologically confirmed prostate cancer through Gleason probability maps. Singh et al. ^7^ established the Histo-MRI protocol for biological validation of advanced MRI techniques using a prospective cohort of patients undergoing biopsy or radical prostatectomy. More recently, Yilmaz et al. ^8^ compared radiologist-derived and bp-MRI DL-based tumor volume estimations with those on whole-mount histopathology as the reference standard. They found that the radiologist-derived volume estimations align better with histopathology than with AI-based methods.

The purpose of the current study is to evaluate the correlation between the spatial extent of a prostate cancer lesion, as identified by an MRI-trained Deep Learning detection model, having as a reference the output of a DL model trained on digital histopathology for cancer detection and the histological and radiological lesion annotations. Our unique rad-path correlation study, with a prospective dataset of patients performing a bp-MRI examination before undergoing radical prostatectomy, has been conducted to compare DL detection models. We claim this work drives the need for the definition of a more solid lesion ground truth for training the DL detection models. This is a necessary step before such bp-MRI trained DL-based models can be used to guide targeted biopsies, reduce unnecessary biopsies, and improve patient stratification for the reduction of overtreatment in patients with prostate cancer.

## Methods

### Deep Learning lesion detection in histopathology

We used a ResNet18 architecture ^9^ trained using Python, PyTorch, and torchvision – 100 epochs, learning rate of 1.0e-4, weight decay of 1.0e-5 – for cancer classification on digital histopathology radical prostatectomy quadrants from diagnostic H&E-stained slides after initialisation with pre-trained ImageNet weights. We used a retrospective balanced dataset of clinically significant prostate cancer histopathology tiles annotated by an expert uropathologist with more than 30 years of clinical experience at Champalimaud Foundation clinical centre (256×256 at 10x resolution) with 110,989, 24,660, and 22,618 tiles (142, 34, and 34 patients) for training, testing and validation, respectively. TrivialAugment ^10^, AugMix ^11^ and random augmentations in the HED space, colour space designed for H&E images, were performed. The DL model obtained was then applied to all prostate histopathology slide fragments for cancer detection at the tile level of a dataset prospectively collected at Champalimaud Foundation clinical centre. We stacked individual reconstructed 2D prostate histopathology slides and filtered small, predicted regions while excluding predictions with low-intensity values.

### Deep Learning lesion detection in MRI

We employed two DL models for prostate lesion detection on MRI images, namely on T2-w MRI only and on bp-MRI, consisting of T2-w, high b-value DWI and ADC images, from a previous work ^12^. The T2-w MRI only model was trained and validated on 623 and 110 cases, respectively, using the ProstateAll dataset which combined T2-w series from ProstateX ^13^, Prostate-158 ^14^ and PROSTATE-Net ^15^ datasets. The bp-MRI model was trained and validated on 286 cases and 72 cases from PROSTATE-Net, respectively. Both models were based on a nnUnet architecture ^16^, and the manual lesion segmentations performed by experienced radiologists were used as ground-truth for the automated lesion segmentation. These models were used for inference on the prospective patients’ MRI images to identify cancerous lesions.

### Evaluation of the DL models in a prospective patient cohort

#### A. Data collection

This study was approved by the Ethics Committee of the Champalimaud Foundation clinical centre, and all patients signed informed consent forms. Patients were included in the study if they had histologically confirmed intermediate or high-risk prostate cancer planned to be treated with radical prostatectomy and were older than 18 years old at the time of diagnosis. Exclusion criteria included any contraindications to mpMRI (non-MRI-safe implants, claustrophobia, physical limitations e.g., back pain), neo-adjuvant/concomitant androgen deprivation therapy, transurethral resection of the prostate (TUR-P) performed within 6 months, and the presence of metastatic disease. We collected and anonymized images from 14 consecutive patients treated with Retzius-sparing robot-assisted laparoscopic radical prostatectomy (RS-RARP) using the da Vinci robotic system (Intuitive Surgical, Sunnyvale, CA). These patients underwent a pre-operative bp-MRI examination in a 3T Philips scanner (Ingenia, Philips Healthcare, The Netherlands) equipped with a 32-channel surface coil. bp-MRI imaging consisted of T2-weighted (T2-w), diffusion-weighted imaging (DWI), and apparent diffusion coefficient (ADC). The T2-weighted (T2-w) MRI was acquired with the following parameters: echo time = 105 ms, repetition time = 4000 ms, field of view = 200 x 200 x 90 mm^3^, in-plane resolution = 0.35 x 0.35 mm^2^, slice thickness = 3 mm, slice number = 30, and gap = 0.3 mm. Additionally, for the Diffusion Weighted Imaging (DWI) the following were used: echo time = 90 ms, repetition time = 5400 ms, field of view = 200 x 200 x 90 mm^3^, in-plane resolution = 0.80 x 0.80 mm^2^, slice thickness = 3 mm, slice number = 33, with no gap, and a diffusion b-value of 0 and 1400 s/mm^2^. Prostate cancer lesions were manually segmented by a radiologist with 6 years of experience (P.S.C.) and by a non-expert radiologist with 2 years of experience (M. U.) on the T2-w images and annotated based on PI-RADS v. 2.1, without prior information on biopsy results. A total of 117 histopathology slides from the whole prostate surgical specimen were sliced, facilitated by the usage of 3D-printed patient-specific molds based on the patients’ pre-operative bp-MRI, as shown in Figure 1 (e). We collected clinical data from the radiological, surgical and histopathological reports, including lesion dimensions, location, PI-RADS, nerve sparing status, International Society of Urological Pathology (ISUP) grade, Gleason score, cribriform growth status, perineural invasion status, extra-prostatic extension, resection margins status, seminal vesical invasion status, T and N stages.

**Figure 1.**
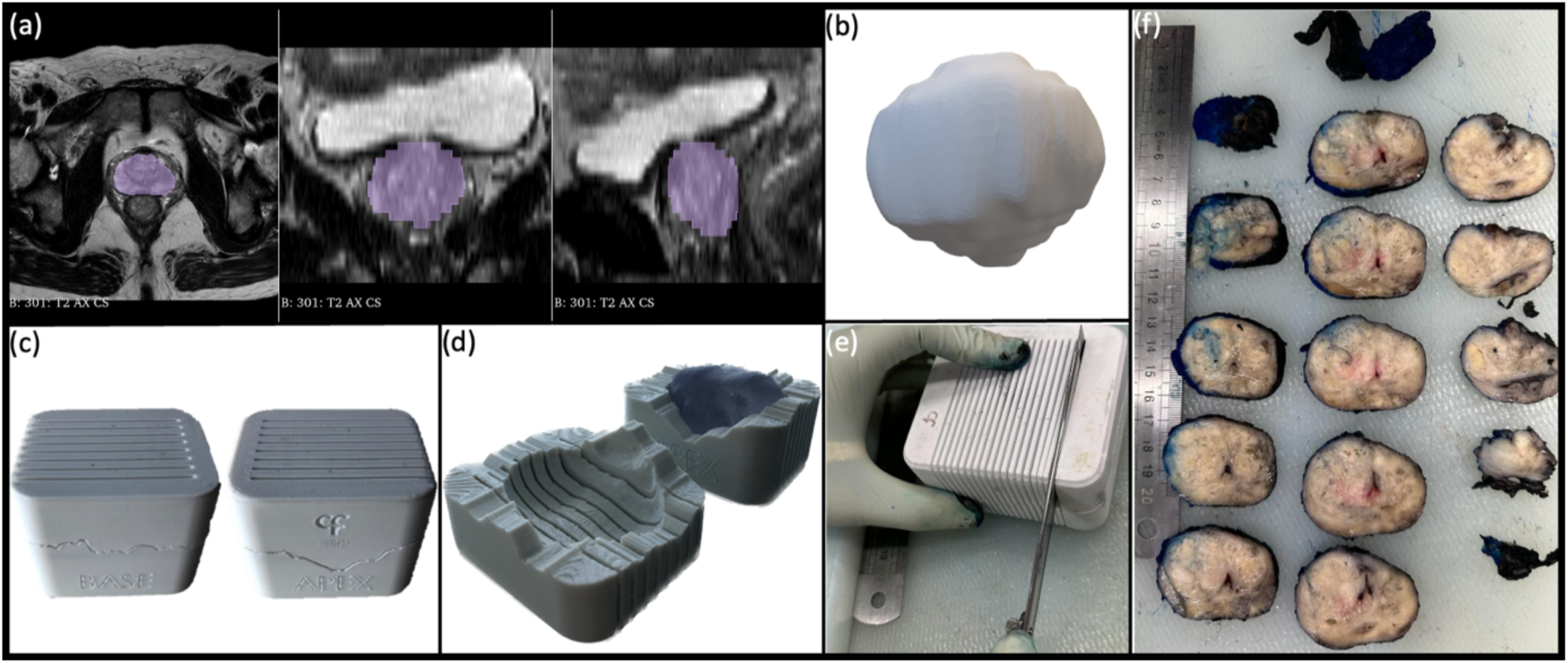
Main prospective data collection steps: (a) validation of the generated automated whole-gland segmentation; (b) creation of the smoothed 3D prostate model using 3D Slicer; (c) and (d) development and 3D printing of the mold, respectively, closed and open views; (e) slicing of the specimen in macroblocks guided by the 3D-printed mold; (f) example of resulting slices spaced by the MRI slice thickness.

#### B. 3D-printing of patient-specific molds

We automatically segmented the prostate gland in the pre-operative T2-w MRI using a DL model from a previous study ^17^. The prostate segmentations were validated, and edited when necessary by a radiologist with 19 years of experience (A.M.G.) – Figure 1 (a). Whole-gland segmentations were exported to standard triangle language (STL) objects in 3D Slicer and smoothed using a median filter with a kernel size of 10 mm to obtain a more polished prostate representation, as shown in Figure 1 (b). The prostate segmentation was smoothed by reducing the number of vertices using Meshmixer (Autodesk, Inc. San Rafael, CA). The resulting model was split in half and the mold was created by extruding two square surfaces into the model using Solidworks (Solidworks, Dassault Systèmes, France). The mold consists of two separate parts that are joined together through a median line automatically calculated from the segmentation, which best encloses the specimen. Additionally, four small pegs were added on both sides. To allow proper prostate placement, the mold was marked with labels for the right (R), base/apex, and anterior (A) prostate sides. A set of 1 mm thick slits were included, spaced by the slice thickness of 3 mm. A margin of 1 mm was set within the mold. The molds were 3D-printed in polylactic acid (PLA) or co-polyester (CPE) filaments using, respectively, Ultimaker 2+ and 3 3D printers (Ultimaker B.V., Utrecht, Netherlands). A prototype of the 3D-printed patient-specific mold is depicted in Figure 1 – (c) and (d).

#### C. Histopathology data preparation

The preservation of the entire prostate specimen was ensured after surgery, and the seminal vesicles were removed. The prostate was placed inside the 3D-printed mold and formalin-fixed at room temperature for at least 24 hours before sectioning. The fixed prostate was sliced, guided by the mold slits, using a single long blade to create 3 mm tissue blocks – Figure 1 – (e) and (f). Larger tissue blocks were cut into halves or quadrants. The standard procedure of pathologic-anatomic diagnosis analysis was conducted on the tissue blocks of 3 mm, including dehydration, paraffin embedding, and microtome sectioning in 3 µm tissue slices starting from the caudal surface. Slides were glass-mounted and stained using haematoxylin and eosin. Slides were digitally scanned in an UltraFast Scanner (Ingenia, Philips Healthcare, The Netherlands) with a resolution of 0.25 × 0.25 µm^2^. The lesions were manually segmented by a pathologist with over 30 years of experience (M.J.B.) using QuPath v.0.4.3 (QuPath developers, The University of Edinburgh, Scotland) ^18^.

#### D. Data pre-processing

The T2-w MRI images were pre-processed, namely, corrected for bias field artifacts using the N4 bias field correction SimpleITK algorithm in Python ^19^ and normalized for image intensities within the range of 0 to 1. The corresponding histopathology slides were identified based on a prostate visual schematic. Out of 117 histopathology slides, 103 were stitched together because the large tissue size prevented whole-mount digitisation. The stitching of histopathology quadrants was done using StitchPro ^20^, a Python-based stitching algorithm developed based on the principles of boundary detection, histogram matching, and differential evolution optimization. The stitching output was quantitatively evaluated by measuring the average Euclidean distance (in millimetres) between corner points of adjacent fragments.

#### E. Rad-path image registration

To improve the 2D co-registration, a binary mask of the prostate was drawn on the T2-w MRI fixed image. Additionally, a set of control points were placed along the prostate boundary at corresponding locations on both images – the T2-w MRI and the histopathology images. These points were automatically generated in the fixed image using the Harris corner detector in Python using scikit-image ^21^. The moving control points were created by scaling and rotating the fixed points to match the corresponding locations in the moving image. After an initial rigid alignment of geometric centres to increase image overlap, an affine transform was performed in Python using itk-elastix ^22^ to register the two masks. This transform was then applied to the corresponding rad-path images. The registration results were visually confirmed and specific cases where misalignment occurred were corrected by rotating or scaling the control points. Our software implementation was evaluated quantitatively using the DSC between the T2-w MRI-based fixed prostate masks and the corresponding histopathology-based resulting image masks. The Python script for rad-path registration can be run through the command line or as a Streamlit application, giving the fixed and moving images, the fixed prostate mask, and the index slice with the number of the corresponding slices to register. This script is available in a GitHub repository.

#### F. Comparison between DL models’ lesion detection outputs and radiological and histopathological annotations

Firstly, we determined the overlap between the bp-MRI model output and the manual radiologists’ lesion annotations, measured by the DSC. We calculated the DSC for each patient between the bp-MRI model output (bpMRI-DL) and Hist-DL, as well as between the Hist-DL and histopathological ground truth (Hist-GT). Lesions were considered detected if there was at least a 10% overlap between the predicted lesion mask and the ground truth to determine true positives (TP), false positives (FP), false negatives (FN), true negatives (TN), and positive predictive value (PPV). We calculated the overlap between each model output and the opposite clinical annotation, where opposite represents Hist-DL vs. Radiologist and bpMRI-DL vs. Hist-GT. All DSC values were compared between each other, using a Kruskal-Wallis test^23^ followed by Dunn’s tests between individual pairs ^24^, corrected for multiple comparisons using the Bonferroni method.

Additionally, we measured the agreement between the two radiologists and between the two MRI DL models (T2-w MRI and bp-MRI) to determine, respectively, if radiologist experience and the input sequence influences the lesions detected.

Finally, we correlated the lesion model output with the tumor characteristics in the radiological and histopathological reports by identifying the presence of detected lesions in the prostate zone identified in the report’s index lesion.

## Results

### Deep Learning lesion detection in histopathology and MRI

The DL model for digital histopathology was evaluated on a retrospective large dataset of histopathology patches annotated at Champalimaud Foundation clinical centre for the presence of clinically significant prostate cancer. The model performed remarkably well for cancer detection with a test set area under the curve (AUC) of 0.99, sensitivity of 0.95, specificity of 0.95, and positive predicted value (PPV) of 0.96.

The nnUNet DL model, trained and tested on T2-w of the ProstateAll dataset, presented fair lesion detection performance. In particular, the model achieved a DSC of 0.34 (95% CI=[0.29, 0.40]) and a sensitivity at 0.1 DSC of 0.64 (95% CI=[0.56, 0.73]). The nnUNet DL model, trained and tested on T2-w, DWI, and ADC images (bp-MRI) of the ProstateAll dataset, presented similar accuracy for lesion segmentation. In particular, the model achieved a DSC of 0.35 (95% CI=[0.29, 0.44]) and sensitivity of 0.63 (95% CI=[0.55, 0.79]).

### Evaluation of the DL models in a prospective clinical-setting dataset

#### A. Stitching and rad-path image registration results

Demographic and clinical data from the radiological, surgical and histopathological reports for the 14 prospective patients are depicted in Table 1. The Python-based stitching solution showed good qualitative and quantitative results. For 103 reconstructed histopathology sections, we obtained a low stitching error, with a mean Euclidean distance between adjacent corner points of 0.30 mm (standard deviation of 0.12 mm). The rad-path registration was conducted on the available slices relevant for the histopathological analysis consisting of a total of 117 slices – corresponding between four and ten slices per patient. Registration results corresponding to multimodal slides are depicted in Figure 2. This process led to a high accuracy on the masks registration (average DSC of 0.90, standard deviation of 0.05). Visual inspection of results showed good overlap between modalities as illustrated in Figure 2, eliminating the need for placing manual points or points inside the structure.

**Figure 2.**
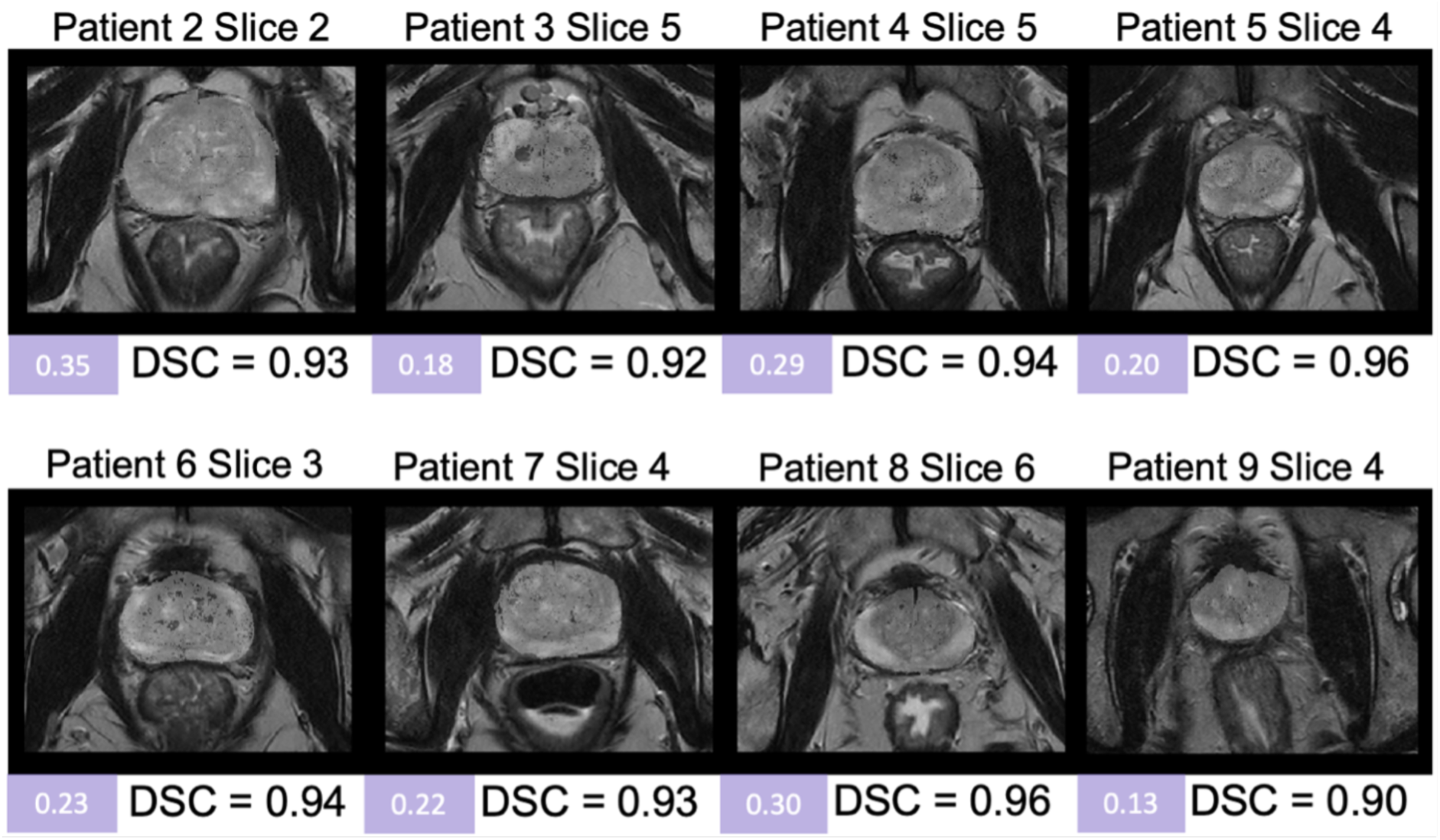
Examples of co-registered histopathology and the corresponding T2-w prostate MRI slice for eight patients. The average Euclidean distance (in mm) from the stitching and the DSC is shown below each registration.

**Table 1.** Prospective patients demographics and clinical data.

| **Characteristic** | **Unit/ Category** | **Median (ICR) / Counts** |
| --- | --- | --- |
| Age | years | 63 [58-67] |
| MRI-surgery interval | days | 78 [34-172] |
| PSA | ng/mL | 8,1 [6,1-9,8] |
| Index lesion maximum diameter | mm | 13 [11-16] |
| Index lesion location | PZ | 11 |
|  | TZ | 1 |
|  | AS | 1 |
|  | TZ+AS | 1 |
| Index lesion PI-RADS | 4 | 9 |
|  | 5 | 5 |
| ISUP | 2 | 10 |
|  | 3 | 3 |
|  | 5 | 1 |
| Gleason score | 3+4 | 10 |
|  | 4+3 | 3 |
|  | 4+5 | 1 |
| Cribiform growth | Present | 5 |
|  | Absent | 9 |
| Perineural invasion | Present | 14 |
| Extraprostatic extension | Present | 5 |
|  | Absent | 8 |
|  | NA | 1 |
| Resection margins | Positive | 6 |
|  | Negative | 8 |
| Seminal vesical invasion | Present | 1 |
|  | Absent | 13 |
| T stage | pT2 | 9 |
|  | pT3a | 4 |
|  | pT3b | 1 |
| N stage | pNx | 8 |
|  | pN0 | 5 |
|  | pN1 | 1 |

#### B. Comparison of lesion extent across modalities and annotations

Inference of DL models was performed on the entire dataset, and Figure 3 shows two representative prostate slices of T2-w and pseudo-whole-mount histopathology with DL model predictions. We can observe that the DL predictions detected in bp-MRI fairly overlap with those detected by the histopathology DL model (Hist-DL). Additionally, we observe no significant lesion detection improvement when adding DWI and ADC to MRI lesion prediction models.

**Figure 3.**
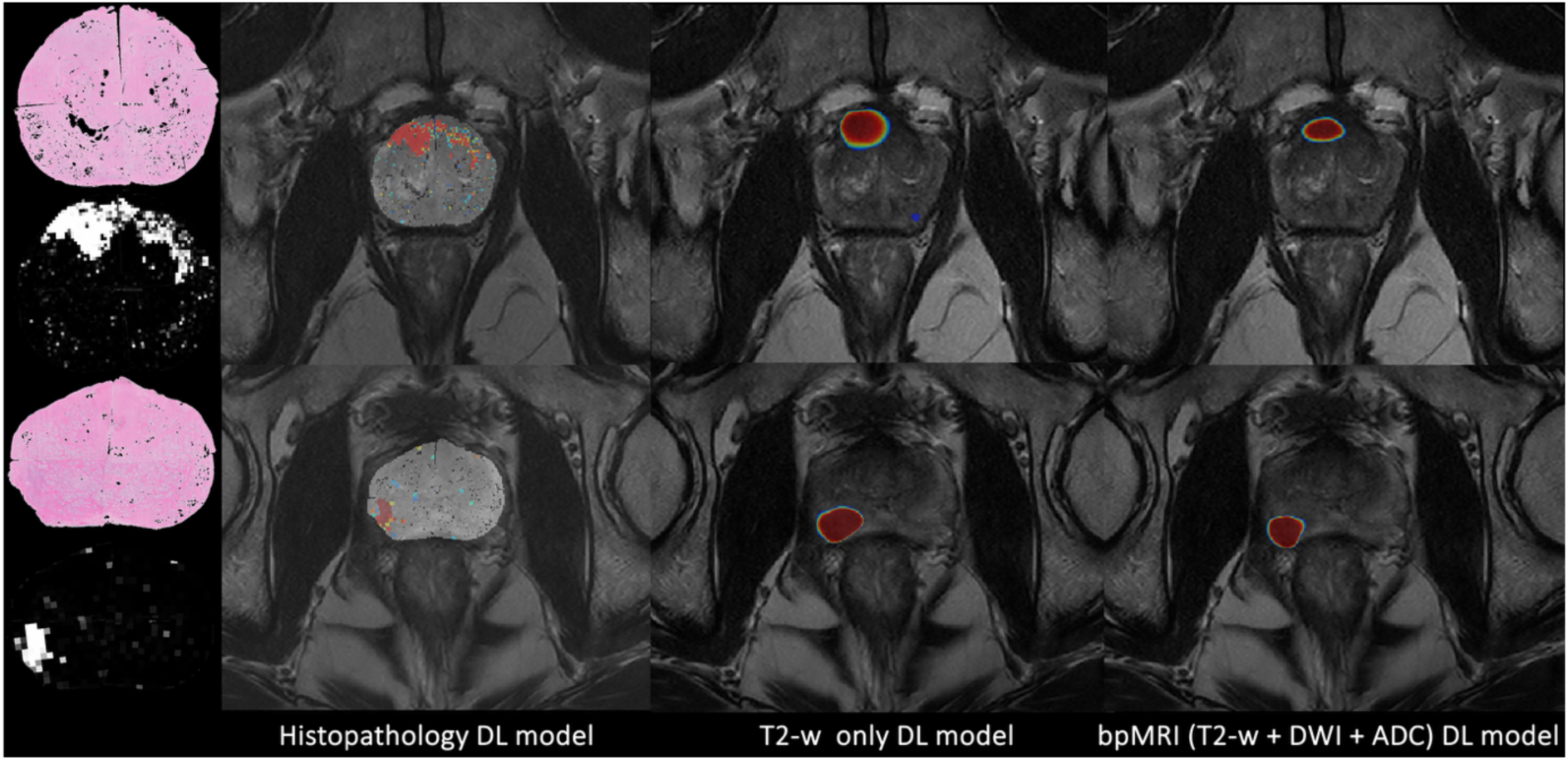
Visual correspondence between rad-path lesion detection models for two patients, in radiological convention. On the left, is the pseudo-whole-mount histopathology section with the DL prediction below (Hist-DL) before filtering small lesion areas. In the second column, the T2-w MRI with the registered histopathology slide and the Hist-DL overlaid. In the last two columns, the T2-w MRI with the DL lesion mask predicted on the T2-w MRI only (T2-DL) and on the bp-MRI (bpMRI-DL), respectively, overlaid.

An average DSC of 0.65 was observed between radiologists on a patient basis, showing good agreement between radiologists. Moreover, the overlap between bpMRI-DL and the radiologists was equivalent for both the experienced radiologist and the non-expert radiologist (DSC of 0.40 and 0.43, respectively). The results of these comparisons on a lesion basis are shown in Figure 4. The average overlap between the experienced radiologist and the Hist-DL was low, with a DSC of 0.10.

**Figure 4.**
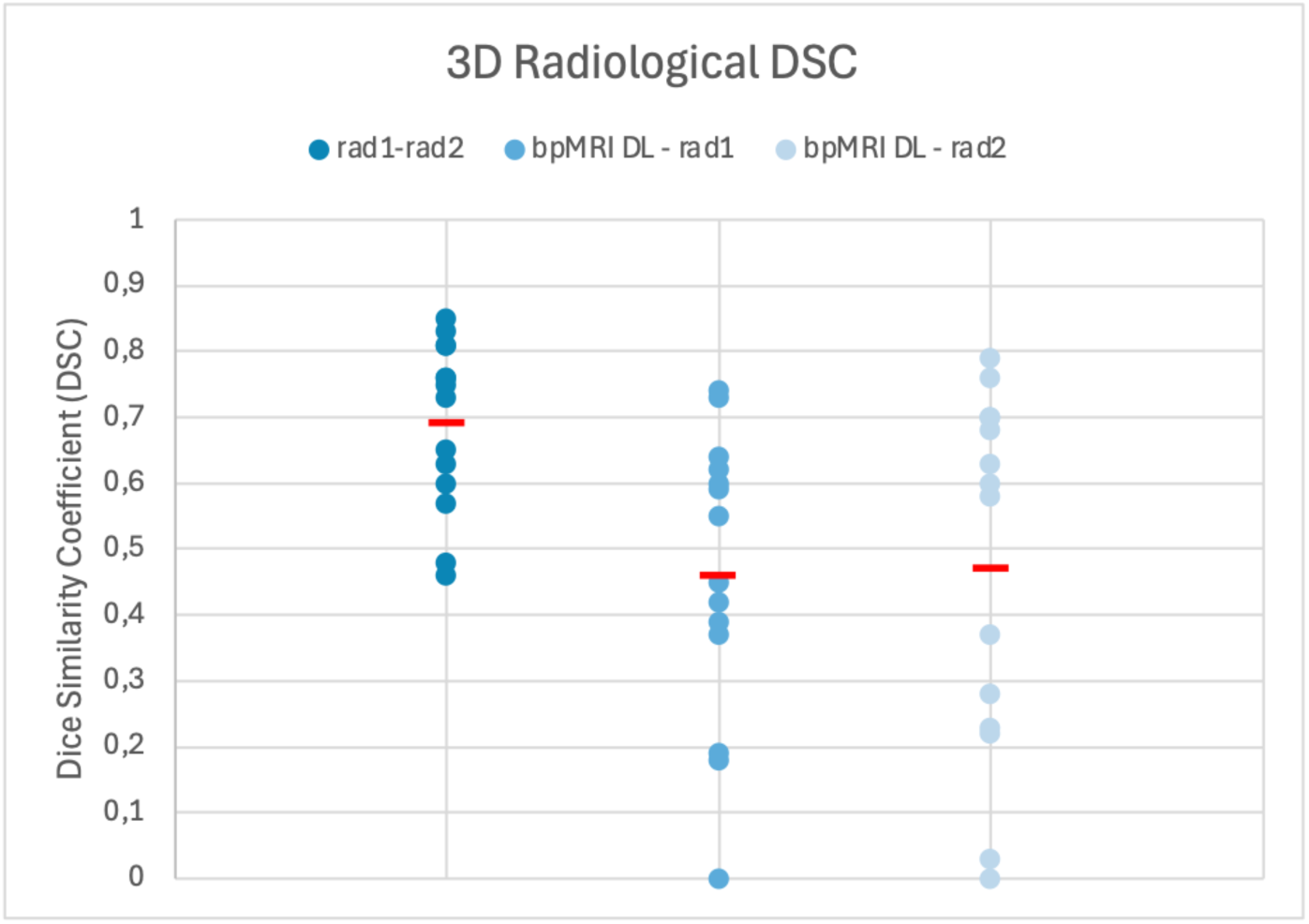
Dice Similarity Coefficient (DSC) of the agreement between radiologists (rad1-rad2) and between radiologists and the bi-parametric MRI (bpMRI) Deep Learning model (bpMRI-DL – rad 1 and bpMRI-DL – rad2) on a lesion basis. Rad1 represents the experienced radiologist, while rad2 is the non-expert radiologist.

Visual comparison between the models and the radiological ground truth for a set of representative patients is shown in Figure 5, showing that some model predictions fairly overlap with ground truth annotations. Considering lesion as detected if there is at least a 10% overlap between prediction and ground-truth, there is a 43 % TP rate (TPR) between the predictions of the bpMRI-DL and Hist-DL models and a 93% TPR between Hist-DL predictions and Hist-GT. Confusion matrices for these comparisons are shown in Figure 6. These comparisons reflect PPVs of 0.46 and 0.93, respectively. Visual comparison of four representative patients of 3D stacked Hist-GT annotations (in blue) and Hist-DL output (in red) is shown in Figure 7.

**Figure 5.**
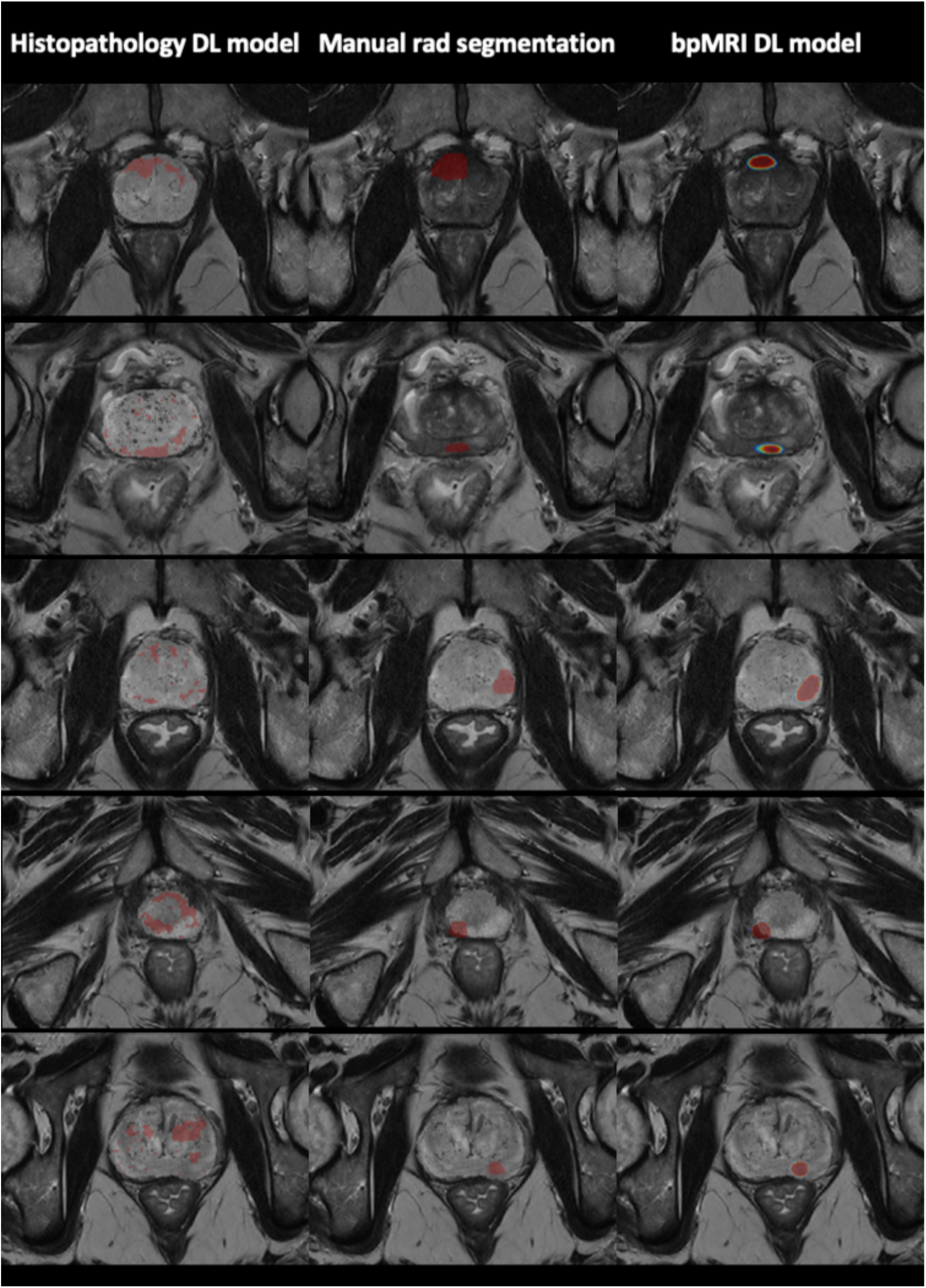
Illustration example of malignant regions in five patients, in radiological convention. On the left column, the histopathology image overlays the corresponding slice of the T2-w MRI image, and the color mask identifies the tumor inferred by the histopathology-based DL model (Hist-DL); in the middle, the color mask identifies the tumor segmented by the radiologist; and on the right the color mask identifies the tumor inferred by the bpMRI-based DL model.

**Figure 6.**
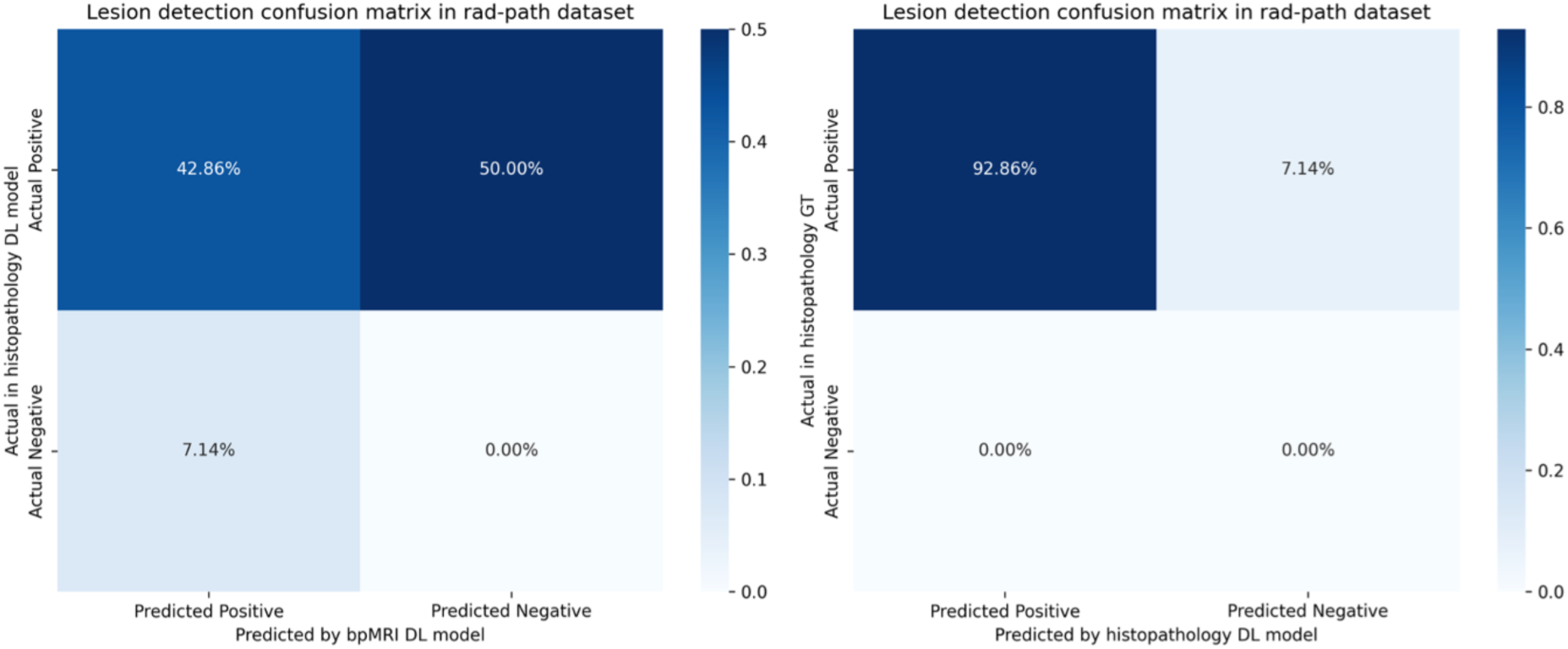
Confusion matrices for overlap above 10%. On the left is the comparison between bpMRI and Histopathology DL model (Hist-DL), while on the right is the comparison between histopathology DL model (Hist-DL) and histopathology ground-truth (Hist-GT).

**Figure 7.**
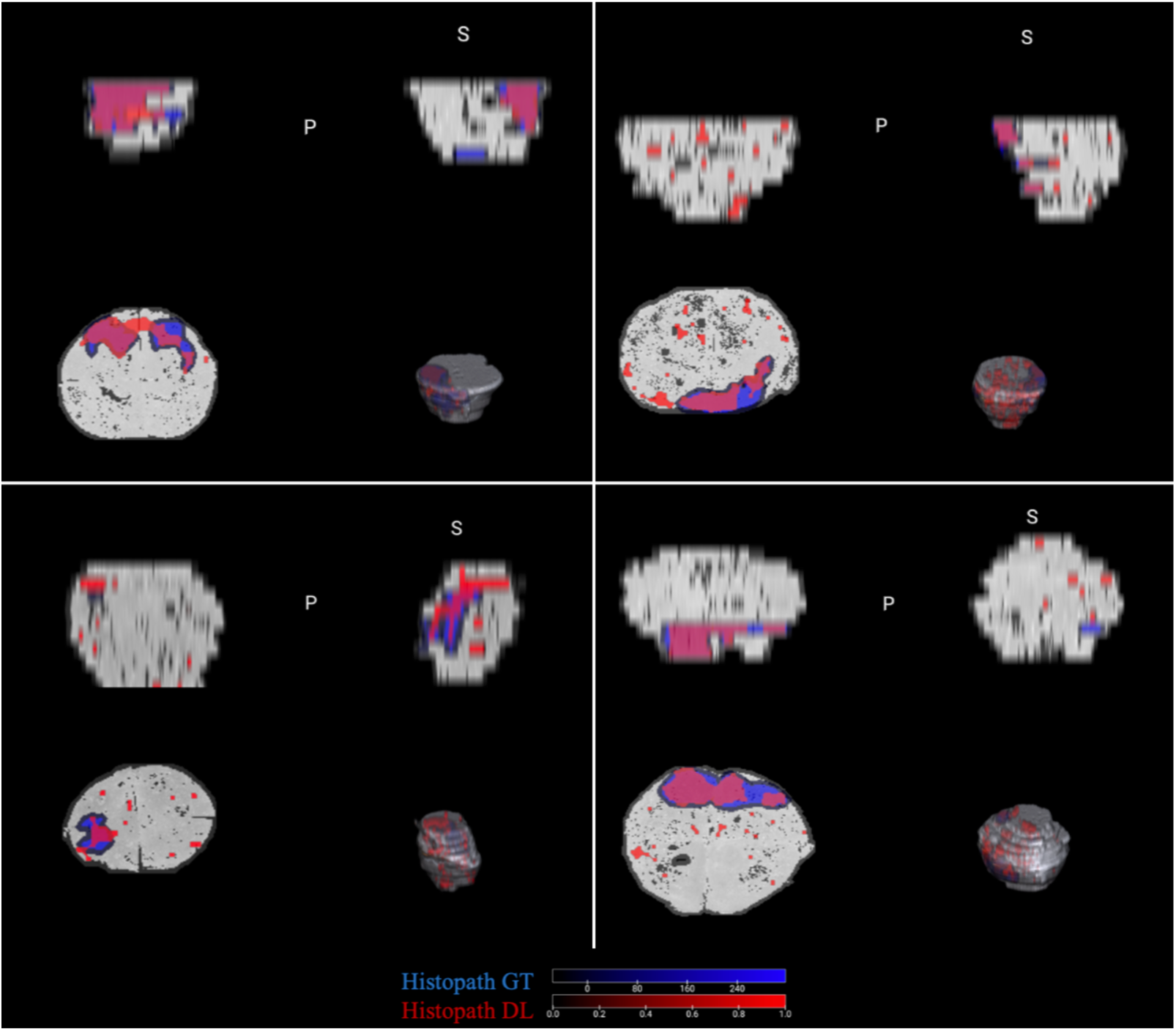
3D stacked histopathology lesion ground-truth annotations overlaid with histopathology DL lesion outputs (Hist-DL) for four representative patients.

Table 2 shows the average DSC of all comparisons on a patient basis. As expected, there is a fair overlap between each DL model and its corresponding annotations (* in Table 2). Conversely, there is a little overlap when comparing both DL models († in Table 2). However, while there were statistically significant differences between comparisons (Kruskal-Wallis chi-squared = 38.26, p-value = 3.3e-7), comparisons between Hist-GT and radiologist annotations were not different from comparisons between Hist-DL and bpMRI-DL annotations (p=1.00 for post-hoc test). Additionally, comparisons between Hist-GT and Hist-DL annotations were not different from comparisons between radiologist and bpMRI-DL annotations (p=1.00 for post-hoc test). Finally, comparisons between Hist-DL and bpMRI-DL annotations were not different from comparisons between Hist-GT and bpMRI-DL (p=1.00 for post-hoc test), or from comparisons between radiologist and Hist-DL (p=1.00 for post-hoc test). Finally, there is a correspondence between the lesion detection model output and the radiological and histopathological report information (Table 3).

**Table 2.** Comparison of DSC scores on a patient basis. This comparison includes the annotations from the radiologist and pathologist (Hist-GT) and the Deep Learning model outputs from bi-parametric MRI (bpMRI-DL) and histopathology (Hist-DL). Lower overlaps are marked with † and higher overlaps are marked with *.

|  |  |  |  |  |
| --- | --- | --- | --- | --- |
| Radiologist | 1.00 | 0.08† | 0.40* | 0.10† |
| Hist-GT |  | 1.00 | 0.09† | 0.41* |
| bpMRI-DL |  |  | 1.00 | 0.10† |
| Hist-DL |  |  |  | 1.00 |
|  | Radiologist | Hist-GT | bpMRI-DL | Hist-DL |

**Table 3.** Radiological characteristics (including maximum diameter of the lesion in the longest axis, lesion location and PI-RADS v2.1) and histopathological characteristics (including International Society of Urological Pathology grade, Gleason score, cribriform growth status, perineural invasion status, extraprostatic extension, resection margins status, seminal vesical invasion status, T and N stages) for the patients described in Figure 5.

| **Characteristic** | **Patient 1** | **Patient 3** | **Patient 4** | **Patient 5** | **Patient 16** |
| --- | --- | --- | --- | --- | --- |
| Maximum diameter | 21 | 13 | 18 | 11 | 8 |
| Location | MRTZa, MRAS | MRPZp, MLPZp, BRPZp, BLPZp | ALPZpl, MLPZpl | ARPZpm, ARPZpl | MLPZpm |
| PI-RADS | 5 | 4 | 5 | 4 | 4 |
| ISUP | 2 | 3 | 2 | 2 | 3 |
| Gleason score | 3+4 | 4+3 | 3+4 | 3+4 | 4+3 |
| Cribriform growth | Absent | Present | Absent | Absent | Absent |
| Perineural invasion | Present | Present | Present | Present | Present |
| Extraprostatic extension | Absent | Present | Present | Absent | Present |
| Resection margins | Positive (R1) | Positive E (R1) | Positive (R1) | Positive E (R1) | Negative (R0) |
| Seminal vesical invasion | Absent | Present | Absent | Absent | Absent |
| T stage | pT2 | pT3b | pT3a | pT2 | pT3a |
| N stage | pN0 | pN1 | pN0 | pNx | pN0 |

## Discussion

In this work, we presented a rad-path correlation approach to compare two DL-based lesion detection models in bp-MRI and histopathology. It was possible to observe this correlation due to the prospective collection of radical prostatectomy surgical specimens histology with corresponding diagnostic prostate bp-MRI. Each surgical specimen was sliced using an in-house developed personalized 3D-printed prostate mold which allowed for accurate matching of slices between histopathology and bp-MRI. Rad-path registration showed good qualitative and quantitative results and the automatic assignment of lesions in both histopathology and prostate cancer demonstrated fair performance, with the automatic bp-MRI model capturing some of the cancer areas that were predicted on histopathology. DL-based bp-MRI lesion predictions showed lower average overlap with histopathology lesion annotations than compared with radiologist lesion annotations. This is expected since the bp-MRI lesion detection DL model was trained with manual radiologists’ segmentations. Both DL-based lesion predictions were consistent with the information present in the radiological and histopathological reports.

Most of the existing studies in rad-path registration have shown relatively high DSC ^25–30^ or low target registration errors below slice thickness ^25,27,31–34^. Our registration solution was a crucial step for rad-path correlation and showed good qualitative and quantitative results compared to existing literature ^33,35,36^ while being an efficient Python-based solution that can be run from the command line or as a Streamlit application. In this paper, we observed a good visual alignment between the histopathology patients’ slides with the corresponding fixed T2-w MRI slice. Also, an average high DSC score of 0.90 reflects a good overlap between the fixed image mask and the registered histopathology slide, representing successful registration results. One important recommendation to guide registration ^37^ is the use of 3D-printed molds to slice the tissue specimens in parallel planes. To optimize tissue sectioning and allow direct slice correspondence, Starobinets et al. developed a mesh basket mold ^25^ while Kalavagunta et al. constructed a sectioning device ^27^. Reynolds et al. ^35^ designed a customized sectioning device with slicing slits placed every five millimetres to eliminate the need for block-face photographs. Wu et al. were the first to combine these molds with *ex vivo* MRI for rad-path registration ^33^. They made changes to a mold designed by Priester et al ^38^ to optimize *ex vivo* MRI. More recently, Sandgren et al. ^34^ also created patient-specific 3D-printed molds, consisting of a cubical shape with two interlocking parts – separated by 11 millimetric thick slits to guide the slicing –, from which the segmented prostate shape from *in vivo* MRI was subtracted. Our 3D-printed patient-specific molds were based on the existing solutions while adding a median line to improve specimen enclosure during slicing.

Although rad-path correlation is limited to prospective studies and is challenging when handling spatial heterogeneity, imaging characteristics have a recognized macroscopic biological association with histopathology data ^39^. Partial or radical resection of organs with cancer allows the direct correspondence between the pre-operative imaging and the tissue histology. Developing 3D-printed patient-specific molds for slicing the organ or lesion of interest according to the preoperative imaging has been proposed in several tumor sites – including brain ^40^, ovary ^41^, kidney ^42^, and prostate ^4^. However, to the best of our knowledge, this is the first time a prospective study carried on operated patients (radical prostatectomies) with prostate cancer is used specifically for the comparison of the lesion extent detected using DL models in bp-MRI and histopathology.

### Limitations

The main limitations of our study include the small sample size, the patient-selection bias (only patients which had indication for radical prostatectomy were included) and the artifacts during tissue processing. The former relates to the time-consuming data collection workflow that relies on the surgical demand. Secondly, although patients undergoing radical prostatectomy offer a unique opportunity to have access to the entire specimen, tumors were confined to the gland not reflecting the entire disease stage variability. Thirdly, are the alterations in the prostate orientation after surgical resection, which compromise the direct comparison between the anatomy of the gland pre and post-operatively. Moreover, during the preparation of histopathology samples and digitisation, tissue loss and artifacts may compromise the stitching results. The latter can be mitigated using a whole-mount scanner for digitisation and promoting quality control on a large dataset.

### Conclusion

In conclusion, we performed a rad-path correlation study between aligned datasets of histopathology and bp-MRI to investigate the level of agreement of the detected lesion’s spatial extent. Both DL MRI-based models and radiologically defined masks present a weak correlation with the ground truth from histopathology, being able to broadly point to the areas where lesions were identified in histopathology. Training AI models with bp-MRI using radiologically defined masks can be problematic given the lack of a high correlation between radiologists and the ground truth from histopathology diagnosis. Better integration of tissue information can potentially improve lesion detection in MRI, with the end goal of enhancing targeted biopsies for better patient stratification.

## Data Availability

The rad-path registration algorithm mentioned in this study is available at a GitHub repository.

https://github.com/CCIG-Champalimaud/RadPath-app

## Acknowledgements

We would like to acknowledge Dr António Beltran for the anatomical pathology histopathological diagnosis (including Gleason scoring and grading), the Anatomical Pathology Service technicians for gross macroscopy tissue preparation and slide digitisation, the Surgical and Nursing teams of the Urology Unit for conducting the radical prostatectomies and nursing care, the Radiology technicians, and Dr Francisco Oliveira for providing feedback on the data analysis. Last but not least, we would like to acknowledge the patients with prostate cancer who agreed in participating in the study.

## References

1 Cornford P, van den Bergh RCN, Briers E, et al. EAU-EANM-ESTRO-ESUR-ISUP-SIOG Guidelines on Prostate Cancer—2024 Update. Part I: Screening, Diagnosis, and Local Treatment with Curative Intent. Eur Urol. 2024;86(2):148–163. doi:10.1016/J.EURURO.2024.03.027

2 Tomaszewski MR, Gillies RJ. The biological meaning of radiomic features. Radiology. 2021;298(3):505–516. doi:10.1148/radiol.2021202553

3 van Houdt PJ, Ghobadi G, Schoots IG, et al. Histopathological Features of MRI-Invisible Regions of Prostate Cancer Lesions. J Magn Reson Imaging. 2020;51(4):1235–1246. doi:10.1002/JMRI.26933

4 Turkbey B, Mani H, Shah V, et al. Multiparametric 3T Prostate Magnetic Resonance Imaging to Detect Cancer: Histopathological Correlation Using Prostatectomy Specimens Processed in Customized Magnetic Resonance Imaging Based Molds. J Urol. 2011;186(5):1818–1824. doi:10.1016/J.JURO.2011.07.013

5 McGarry SD, Hurrell SL, Iczkowski KA, et al. Radio-pathomic Maps of Epithelium and Lumen Density Predict the Location of High-Grade Prostate Cancer. Int J Radiat Oncol Biol Phys. 2018;101(5):1179–1187. doi:10.1016/j.ijrobp.2018.04.044

6 McGarry SD, Bukowy JD, Iczkowski KA, et al. Gleason probability maps: A radiomics tool for mapping prostate cancer likelihood in MRI space. Tomography. 2019;5(1):127–134. doi:10.18383/j.tom.2018.00033

7 Singh S, Mathew M, Mertzanidou T, et al. Histo-MRI map study protocol: a prospective cohort study mapping MRI to histology for biomarker validation and prediction of prostate cancer. BMJ Open. 2022;12(4):e059847. doi:10.1136/bmjopen-2021-059847

8 Yilmaz EC, Harmon SA, Lis RT, et al. Evaluating deep learning and radiologist performance in volumetric prostate cancer analysis with biparametric MRI and histopathologically mapped slides. Abdominal Radiology 2024. Published online December 11, 2024:1–13. doi:10.1007/S00261-024-04734-6

9 He K, Zhang X, Ren S, Sun J. Deep Residual Learning for Image Recognition. Proceedings of the IEEE Computer Society Conference on Computer Vision and Pattern Recognition. 2015;2016-December:770–778. doi:10.1109/CVPR.2016.90

10 Müller SG, Hutter F. TrivialAugment: Tuning-free Yet State-of-the-Art Data Augmentation. Proceedings of the IEEE International Conference on Computer Vision. Published online March 18, 2021:754–762. doi:10.1109/ICCV48922.2021.00081

11 Hendrycks D, Mu N, Cubuk ED, Zoph B, Gilmer J, Lakshminarayanan B. AugMix: A Simple Data Processing Method to Improve Robustness and Uncertainty. 8th International Conference on Learning Representations, ICLR 2020. Published online December 5, 2019. Accessed December 20, 2024. https://arxiv.org/abs/1912.02781v2

12 Rodrigues NM, Almeida JG de, Verde ASC, et al. Analysis of domain shift in whole prostate gland, zonal and lesions segmentation and detection, using multicentric retrospective data. Comput Biol Med. 2024;171. doi:10.1016/J.COMPBIOMED.2024.108216

13 PROSTATEX - The Cancer Imaging Archive (TCIA). Accessed December 4, 2024. https://www.cancerimagingarchive.net/collection/prostatex/

14 Adams LC, Makowski MR, Engel G, et al. Prostate158 - An expert-annotated 3T MRI dataset and algorithm for prostate cancer detection. Comput Biol Med. 2022;148:105817. doi:10.1016/J.COMPBIOMED.2022.105817

15 ProstateNET | Prostate Imaging Archive. Accessed December 4, 2024. https://prostatenet.eu/

16 Isensee F, Jaeger PF, Kohl SAA, Petersen J, Maier-Hein KH. nnU-Net: a self-configuring method for deep learning-based biomedical image segmentation. Nat Methods. 2021;18(2):203–211. doi:10.1038/S41592-020-01008-Z

17 Rodrigues NM, Silva S, Vanneschi L, Papanikolaou N. A Comparative Study of Automated Deep Learning Segmentation Models for Prostate MRI. Cancers *2023, Vol 15, Page* 1467. 2023;15(5):1467. doi:10.3390/CANCERS15051467

18 Bankhead P, Loughrey MB, Fernández JA, et al. QuPath: Open source software for digital pathology image analysis. Scientific Reports *2017 7:1.* 2017;7(1):1–7. doi:10.1038/s41598-017-17204-5

19 Tustison NJ, Avants BB, Cook PA, et al. N4ITK: improved N3 bias correction. IEEE Trans Med Imaging. 2010;29(6):1310–1320. doi:10.1109/TMI.2010.2046908

20 Verde ASC, De Almeida JG, Fonseca J, Matos C, Conceição RC, Papanikolaou N. StitchPro for Computational Pathology Stitching in Patients with Prostate Cancer. Proceedings - International Symposium on Biomedical Imaging. Published online 2024. doi:10.1109/ISBI56570.2024.10635793

21 Van Der Walt S, Schönberger JL, Nunez-Iglesias J, et al. Scikit-image: Image processing in python. PeerJ. 2014;2014(1):e453. doi:10.7717/PEERJ.453/FIG-5

22 Klein S, Staring M, Murphy K, Viergever MA, Pluim JPW. elastix: a toolbox for intensity-based medical image registration. IEEE Trans Med Imaging. 2010;29(1):196–205. doi:10.1109/TMI.2009.2035616

23 Kruskal WH, Wallis WA. Use of Ranks in One-Criterion Variance Analysis. J Am Stat Assoc. 1952;47(260):583–621. doi:10.1080/01621459.1952.10483441

24 Dunn OJ. Multiple Comparisons Using Rank Sums. Technometrics. 1964;6(3):241–252. doi:10.1080/00401706.1964.10490181

25 Starobinets O, Guo R, Simko JP, et al. Semiautomatic registration of digital histopathology images to in vivo MR images in molded and unmolded prostates. J Magn Reson Imaging. 2014;39(5):1223–1229. doi:10.1002/JMRI.24287

26 Li L, Pahwa S, Penzias G, et al. Co-Registration of ex vivo Surgical Histopathology and in vivo T2 weighted MRI of the Prostate via multi-scale spectral embedding representation. Sci Rep. 2017;7(1). doi:10.1038/S41598-017-08969-W

27 Kalavagunta C, Zhou X, Schmechel SC, Metzger GJ. Registration of in vivo prostate MRI and pseudo-whole mount histology using Local Affine Transformations guided by Internal Structures (LATIS). Journal of Magnetic Resonance Imaging. 2015;41(4):1104–1114. doi:10.1002/jmri.24629

28 Shao W, Banh L, Kunder CA, et al. ProsRegNet: A deep learning framework for registration of MRI and histopathology images of the prostate. Med Image Anal. 2020;68:101919. doi:10.1016/j.media.2020.101919

29 Rusu M, Shao W, Kunder CA, et al. Registration of presurgical MRI and histopathology images from radical prostatectomy via RAPSODI. Med Phys. 2020;47(9):4177–4188. doi:10.1002/mp.14337

30 Sood RR, Shao W, Kunder C, et al. 3D Registration of pre-surgical prostate MRI and histopathology images via super-resolution volume reconstruction. Med Image Anal. 2021;69:101957. doi:10.1016/j.media.2021.101957

31 Ward AD, Crukley C, Mckenzie CA, et al. Prostate: Registration of Digital Histopathologic Images to in Vivo MR Images Acquired by Using Endorectal Receive Coil. 2012;263(3). doi:10.1148/radiol.12102294/-/DC1

32 Orczyk C, Rusinek H, Rosenkrantz AB, et al. Preliminary experience with a novel method of three-dimensional co-registration of prostate cancer digital histology and in vivo multiparametric MRI. Clin Radiol. 2013;68(12). doi:10.1016/J.CRAD.2013.07.010

33 Wu HH, Priester A, Khoshnoodi P, et al. A system using patient-specific 3D-printed molds to spatially align in vivo MRI with ex vivo MRI and whole-mount histopathology for prostate cancer research. Journal of Magnetic Resonance Imaging. 2019;49(1):270–279. doi:10.1002/jmri.26189

34 Sandgren K, Nilsson E, Keeratijarut Lindberg A, et al. Registration of histopathology to magnetic resonance imaging of prostate cancer. Phys Imaging Radiat Oncol. 2021;18:19–25. doi:10.1016/J.PHRO.2021.03.004

35 Reynolds HM, Williams S, Zhang A, et al. Development of a registration framework to validate MRI with histology for prostate focal therapy. Med Phys. 2015;42(12):7078–7089. doi:10.1118/1.4935343

36 Losnegård A, Reisæter L, Halvorsen OJ, et al. Intensity-based volumetric registration of magnetic resonance images and whole-mount sections of the prostate. Computerized Medical Imaging and Graphics. 2018;63:24–30. doi:10.1016/j.compmedimag.2017.12.002

37 Alyami W, Kyme A, Bourne R. Histological Validation of MRI: A Review of Challenges in Registration of Imaging and Whole-Mount Histopathology. Journal of Magnetic Resonance Imaging. 2022;55(1):11–22. doi:10.1002/JMRI.27409

38 Priester A, Natarajan S, Khoshnoodi P, et al. Magnetic Resonance Imaging Underestimation of Prostate Cancer Geometry: Use of Patient Specific Molds to Correlate Images with Whole Mount Pathology. J Urol. 2017;197(2):320–326. doi:10.1016/J.JURO.2016.07.084

39 Park JE, Park SY, Kim HJ, Kim HS. Reproducibility and Generalizability in Radiomics Modeling: Possible Strategies in Radiologic and Statistical Perspectives. Korean J Radiol. 2019;20(7):1124. doi:10.3348/kjr.2018.0070

40 Bobholz SA, Lowman AK, Barrington A, et al. Radiomic Features of Multiparametric MRI Present Stable Associations With Analogous Histological Features in Patients With Brain Cancer. Tomography. 2020;6(2):160. doi:10.18383/J.TOM.2019.00029

41 Delgado-Ortet M, Reinius MA V., McCague C, et al. Lesion-specific 3D-printed moulds for image-guided tissue multi-sampling of ovarian tumours: A prospective pilot study. Front Oncol. 2023;13. doi:10.3389/fonc.2023.1085874

42 Dwivedi DK, Chatzinoff Y, Zhang Y, et al. Development of a Patient-specific Tumor Mold Using Magnetic Resonance Imaging and 3-Dimensional Printing Technology for Targeted Tissue Procurement and Radiomics Analysis of Renal Masses. Urology. 2018;112:209–214. doi:10.1016/j.urology.2017.08.056

